# Pediatric health system impact of an early respiratory viral season in Eastern Ontario, Canada: A descriptive analysis

**DOI:** 10.1101/2022.12.17.22283614

**Authors:** Nisha Thampi, Lynn Meng, Liam Bruce, Connor McLean, Melanie Buba, Lise Bisnaire, Ken J. Farion, Lindy M. Samson

**Author notes:** **Corresponding author’s email address**. **Funding statement** None. **Declaration of author(s) competing interests** None.

## Abstract

**Background:** The current respiratory viral season in Ontario started early with an intensity experienced throughout the pediatric health system. We sought to examine trends in patient volumes and level of care intensity among children admitted with laboratory-confirmed respiratory viral infection over the last five years in Ottawa.

**Methods:** This was a retrospective cohort study of patients at CHEO, a pediatric health centre in Ottawa, who were diagnosed with a laboratory-confirmed respiratory viral infection in the first 72 hours of admission between October 22, 2017 and December 10, 2022. Their admissions were stratified by age groups and levels of care intensity and evaluated for trends over six surveillance periods that begin in Week 35 and end in Week 34 of the following calendar year.

**Results:** During this current surveillance period, there was an early, rapid, two-fold increase in admissions due to respiratory viral infections compared to previous periods, driven largely by RSV and influenza. While there were similar age distributions, there was a larger volume of Level 2 and 3 admissions, and higher proportion of patients requiring Level 2 intensity of care (20.8% versus 2.2% to 12.0% in pre-pandemic years; p<0.001). Lengths of stay were comparable to pre-pandemic surveillance years.

**Interpretation:** The current viral season has been associated with elevated volumes and higher inpatient acuity compared to previous years and underscores the need for additional operational and human health resources to support pediatric health systems.

## Introduction

The cyclical nature of respiratory syncytial virus (RSV) and influenza activity in temperate climates was disrupted during the COVID-19 pandemic (1). Before March 2020 in Ontario, elevated viral activity would begin in December and continue through the Winter (2). In 2022, the experience in the Southern Hemisphere suggested pediatric health systems in the Northern Hemisphere should anticipate and prepare for earlier and larger-than-typical viral activity (3,4). Predictably, by mid-October, percent positivity for RSV and influenza was above 5% in the community, signifying high circulation, and we began to experience larger-than-expected pediatric demand for care (5).

CHEO is a tertiary pediatric centre in Ottawa serving 500,000 children and youth from Eastern Ontario, Western Quebec, Northern Ontario, and Nunavut. The early surge in RSV activity in October, followed closely by influenza, required the opening of additional pediatric intensive care unit (PICU) and inpatient medicine beds to meet the needs of infants and children with severe illnesses, creating an average critical care occupancy of 152% over the six weeks leading up to this report.

To characterize the relative impact of this viral season on our pediatric health system, we examined trends in volumes and levels of care intensity among children admitted with laboratory-confirmed respiratory viral infection over the last five years.

## Methods

### Study cohort

This is a single-centre, retrospective, descriptive cohort study that examined electronic health record data (Epic Systems Corp, Verona, WI) from patients admitted to CHEO between October 22^nd^, 2017 and December 10th, 2022, inclusive.

The cohort consisted of all inpatients aged 18 years and younger with laboratory-confirmed respiratory viral infection. Patients were included if one or more viral pathogens were detected by polymerase chain reaction (PCR) of a nasopharyngeal swab ordered within the first 72 hours of admission. Respiratory viruses were reported as one of the following: RSV; Influenza A virus; Influenza B virus; severe acute respiratory syndrome coronavirus 2 (SARS-CoV-2); or those that are part of the extended respiratory viral panel (adenovirus; parainfluenza virus 1, 2, 3, and 4; human metapneumovirus, enterovirus/rhinovirus, seasonal coronaviruses).

### Analysis

Patients were categorized by their date of admission, age, type of virus detected at admission, and highest level of care required. When grouped by year of admission, surveillance years were defined by epidemiological week starting on the 35^th^ week of one year (approximately early September) and ending on the 34^th^ week of the following year (end-August) (6). Thus, six surveillance periods were analysed for trends: 2017/18, 2018/19, 2019/20, 2020/21, 2021/22 and the current period, 2022/23. Patient age was defined as the age at admission. Patient level of care intensity groups were mutually exclusive, with Level 3 defined as patients admitted to the pediatric intensive care unit (PICU) or the additional PICU overflow spaces in the surgical day unit and the neonatal intensive care unit (NICU) at any point in a given census time period. Level 2 was defined as patients on inpatient medicine units at CHEO who required advanced respiratory or nursing supports, including receipt of high-flow oxygen, non-invasive positive pressure ventilation and presence of a tracheostomy, but were not admitted to the Level 3 intensive care unit (or PICU overflow spaces) at the time of census. Level 1 was defined as all admitted patients not receiving Level 3 or Level 2 care.

Patient characteristics were summarized using counts and proportions, or medians and quartiles, where appropriate. Differences between patient categories were compared using Kruskal-Wallis tests for continuous variables, and Pearson Chi-square tests for categorical variables. Post-hoc analyses, including Dunn’s Test of Multiple Comparisons with Bonferroni adjustment for continuous variables, and pairwise Chi-square tests with Bonferroni adjustment for categorical variables, were used to compare individual groups.

Admission data over time were summarized via graphical representation by epidemiological week and year of admission, or continuously by date of admission. The week was defined as beginning on Sunday.

The weekly number of inpatients at CHEO stratified by the type of respiratory virus detected at admission was visualized via line graph, with the total number of inpatients at CHEO as a comparison. The number of admissions positive for Influenza (A or B) virus and RSV were stratified by patient admission year. The trends in the number of Level 2 and Level 3 patients admitted with a positive respiratory virus test were stratified by patient age at admission categories and visualized by line graph.

The numeric outputs and figures for this article were generated using R version 4.2.0 (7). Statistical significance levels for all tests were set to *p* < 0.05.

Research ethics review was sought but waived, as the study was part of a quality improvement initiative to urgently inform and adjust hospital operational planning, with secondary use of patient-level data.

## Results

A total of 28,077 children were admitted to CHEO from October 22^nd^, 2017 to December 10^th^, 2022 and were assessed for eligibility (Table 1). Of these, 2,935 (10.5%) had a respiratory virus detected within 72 hours of admission and were included in this cohort. RSV was detected in 1,365 children (46.5%), Influenza A or B in 482 children (16.4%), and SARS-CoV-2 in 292 children (9.9%) during the study period (Table S1).

**Table 1:**
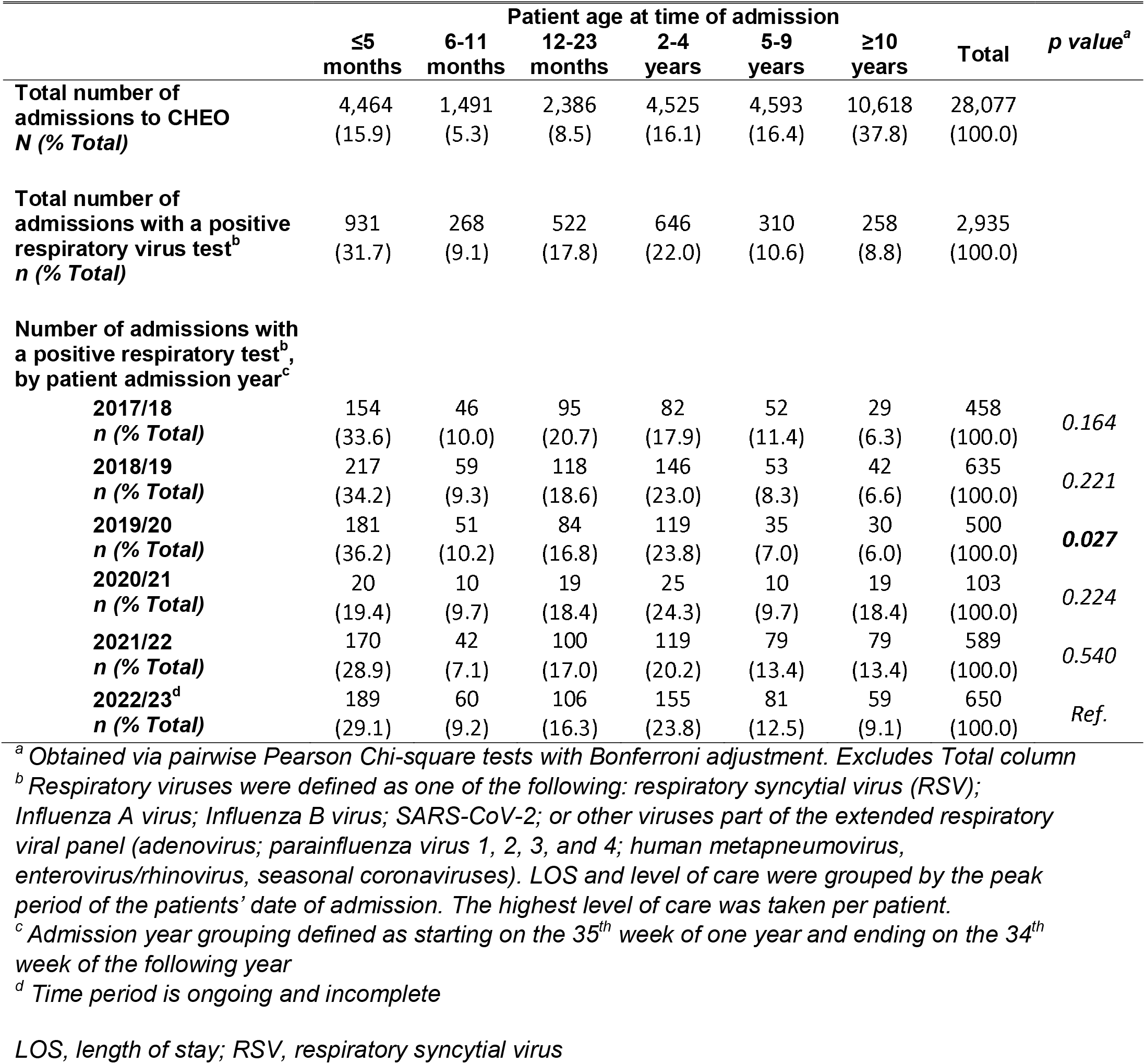
Patient characteristics by patient age at admission.

At the time of writing, weekly admissions for laboratory-confirmed respiratory viral illnesses during the current surveillance year had peaked at 87 during Week 47, during which nearly half of all inpatients at CHEO were admitted with a laboratory-confirmed respiratory viral illness, reflecting a two-fold increase in number and proportion of admissions, compared to pre-pandemic years (Figure 1).

**Figure 1:**
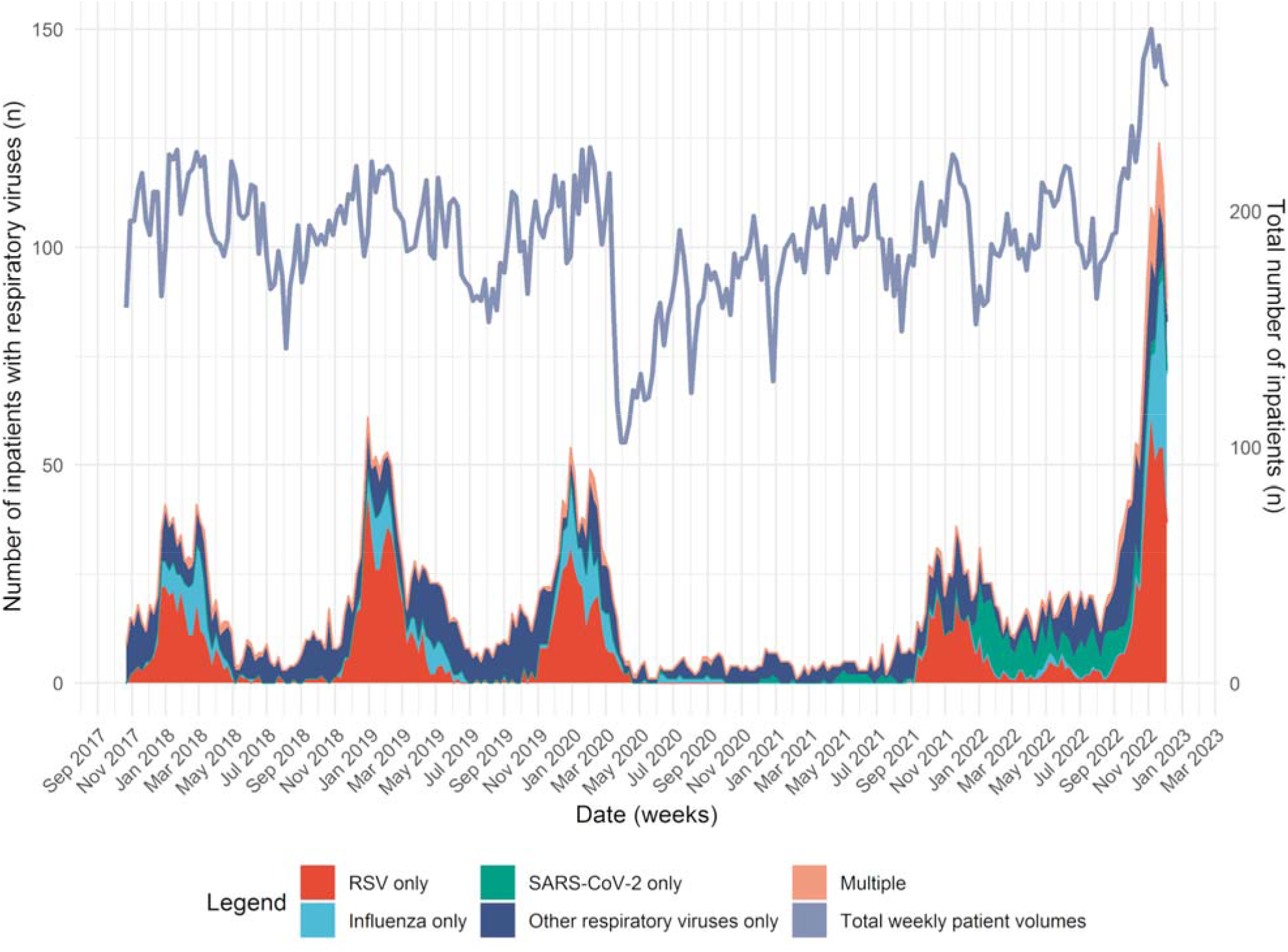
Total number of inpatients and number of inpatients with respiratory viruses detected at admission per week. Primary Y axis corresponds to the stacked areas in plot and shows the weekly number of inpatients at CHEO with a respiratory virus detected at time of admission, stratified by type of virus (categories are mutually exclusive). “Influenza only” group includes one of Influenza A or B viruses. “Other” group includes other viruses that are part of the extended respiratory viral panel (adenovirus; parainfluenza virus 1, 2, 3, and 4; human metapneumovirus, enterovirus/rhinovirus, seasonal coronaviruses). “Multiple” includes any combination of two or more viruses detected at time of admission. Secondary Y axis and line reflect the weekly number of all inpatients at CHEO. *RSV, respiratory syncytial virus; SARS-CoV-2, severe acute respiratory syndrome coronavirus-2*

RSV-related admission volumes began to increase before the start of the 2022/23 surveillance year (Figure 2A), similar to 2021/22, but 10 weeks earlier than pre-pandemic periods, with a 1.5-fold increase in peak number of admissions in one week, compared to the next highest peak in 2018/19. Influenza-related admissions rapidly progressed over the 42^nd^ epidemiological week of 2022/23 (Figure 2B), 4 to 6 weeks earlier than in pre-pandemic periods and with, at the time of writing, a peak weekly number of admissions at the end of November that was 2-fold greater than the next highest peak in 2019/20.

**Figure 2:**
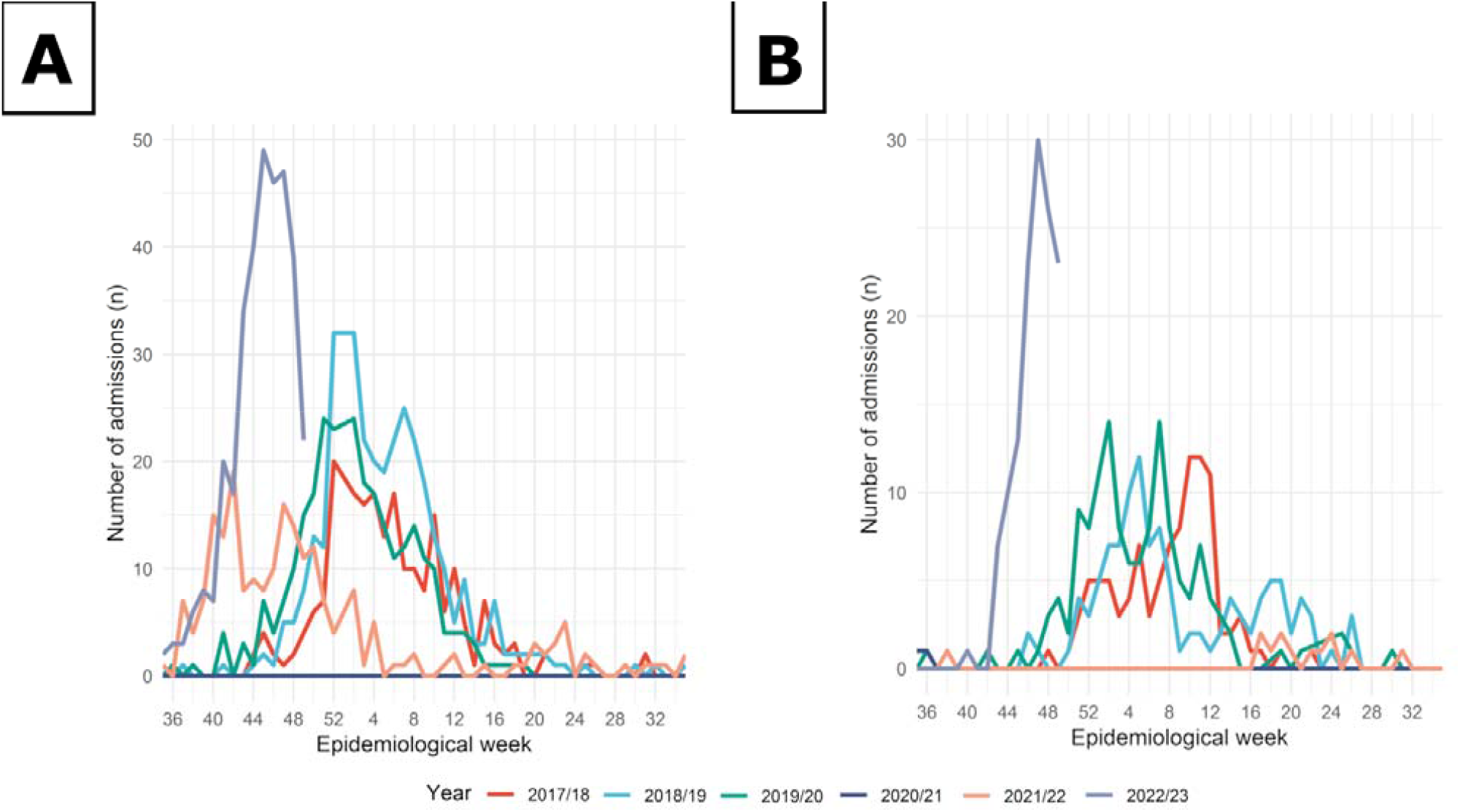
Number of admissions per week with a positive RSV and Influenza test by year of admission groups, October 22, 2017, to December 10, 2022. **(A)** Comparison of number of children admitted to CHEO with a positive test for RSV per week, stratified by year of admission categories. Respiratory samples were collected within 72 hours or more after admission. **(B)** Comparison of the number of children admitted to CHEO with a positive test for Influenza A or B virus per week, stratified by year of admission categories. Positive tests exclude orders collected 72 hours or more after admission. *RSV, respiratory syncytial virus*

In the current period, children under 6 months accounted for the highest proportion (29.1%) of those with positive respiratory tests, followed by children ages 2 to 4 years (23.8%), and 12 to 23 months (16.3%) (Table 1). The age distribution of patients in the 2022/23 cohort were comparable to past years (*p*>0.05), except for an older cohort of patients in 2019/20 (p=0.027).

The distribution of patients by level of care over time is represented as monthly trends stratified by patient age group at admission in Figure 3 and as patient admission year groups in Table 2. During the peak month of viral activity this period (November), there was a 4-fold increase in Level 2 admissions compared to previous years, and a nearly-3-fold increase in Level 3 admissions compared to previous years. Relative to other years and other levels of care, the proportion of Level 2 admissions this year has increased (*p*=0.004 *vs* 2021/22; *p*<0.001 *vs* all other years) (Table 2). Median length of stay (LOS) during the current period has been 3 days (Q1-Q3: 2-5 days), comparable to previous years (*p*>0.05) except for 2020/21 (5 days [Q1-Q3: 3-10 days], *p*<0.001) (Table 2).

**Table 2:**
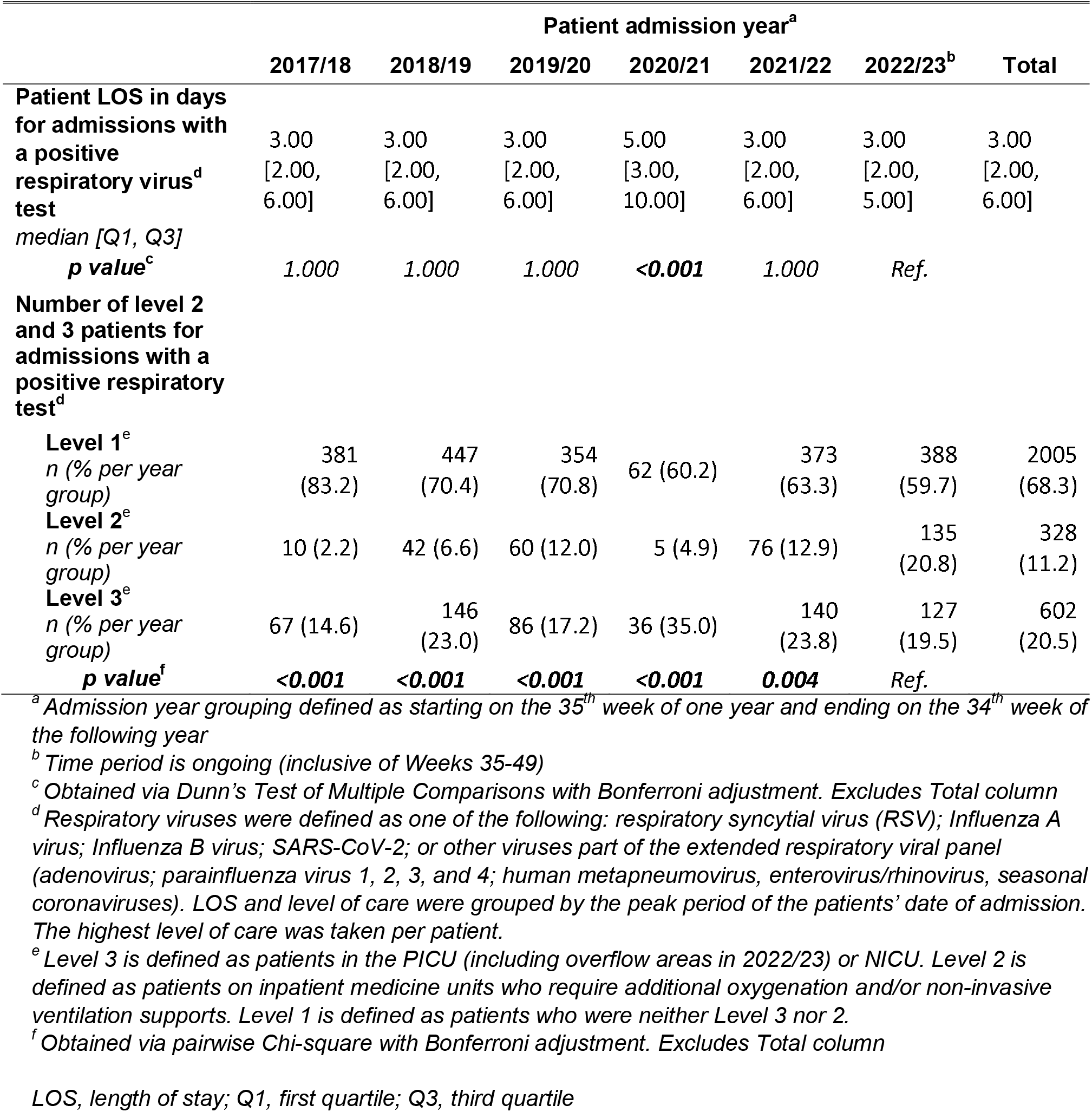
Patient characteristics by patient admission year.

**Figure 3:**
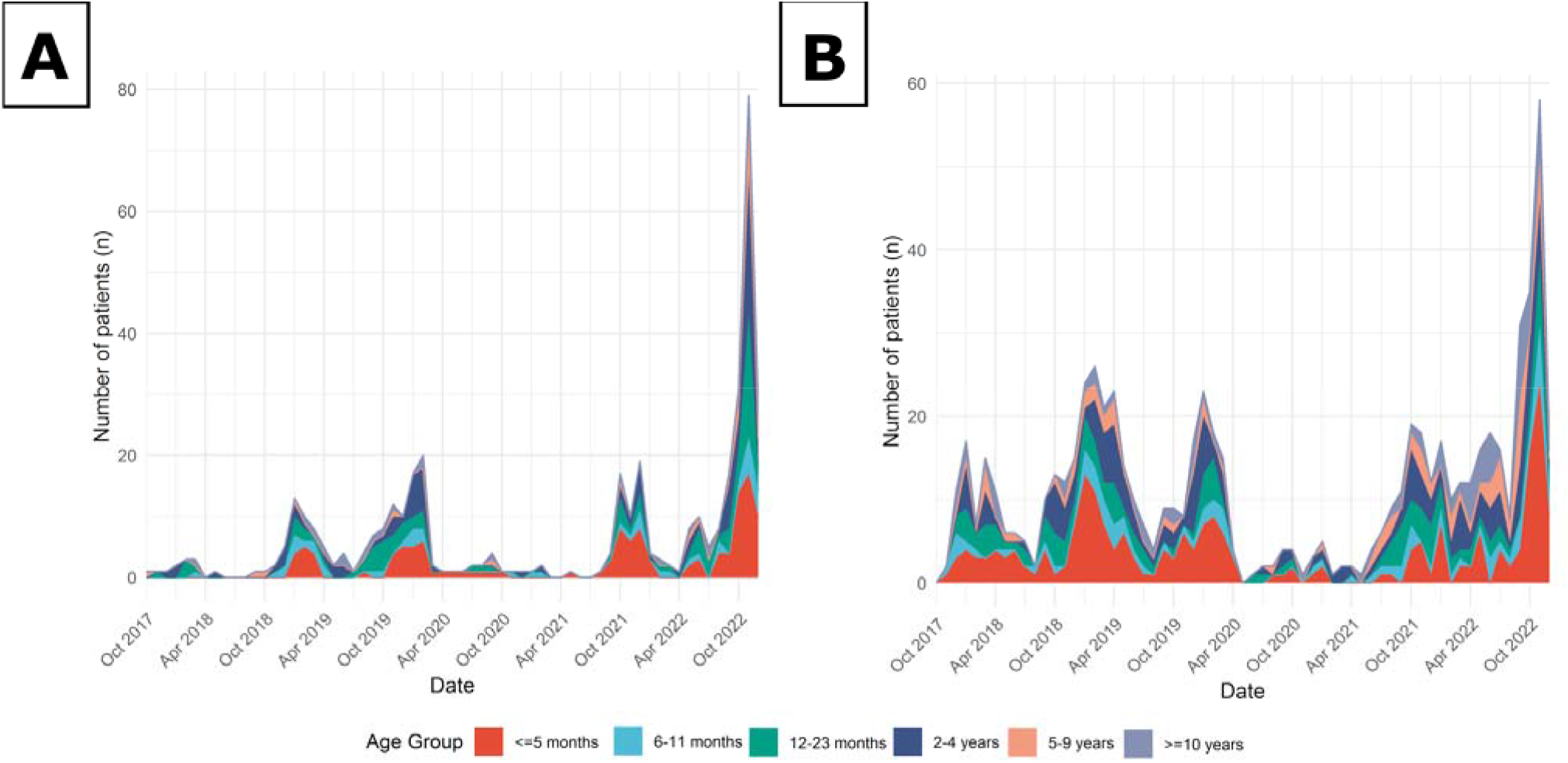
Number of patients by level of care and age per month with a positive respiratory virus test at admission. **(A)** Number of level 2 patients at CHEO with a positive respiratory virus test at time of admission per month by patient age at admission. Level 2 is defined as patients on inpatient medicine units who require high-flow oxygen and/or non-invasive ventilation supports. Respiratory viruses were defined as one of the following: RSV; Influenza A virus; Influenza B virus; SARS-CoV-2; or other viruses in the extended respiratory viral panel (adenovirus; parainfluenza virus 1, 2, 3, and 4; human metapneumovirus, enterovirus/rhinovirus, seasonal coronaviruses). **(B)** Number of level 3 patients at CHEO with a positive respiratory virus test at time of admission per month by patient age at admission. Level 3 is defined as patients in the PICU or NICU. Respiratory viruses were defined as one of the following: RSV; Influenza A virus; Influenza B virus; SARS-CoV-2; or other viruses in the extended respiratory viral panel (adenovirus; parainfluenza virus 1, 2, 3, and 4; human metapneumovirus, enterovirus/rhinovirus, seasonal coronaviruses). *NICU, neonatal intensive care unit; PICU, pediatric intensive care unit; RSV, respiratory syncytial virus; SARS-CoV-2, severe acute respiratory syndrome coronavirus 2*

## Interpretation

For our hospital and the children we serve, the current surveillance period has been associated with an earlier, steeper and two-fold increase in pediatric admissions spanning all levels of care intensity for laboratory-confirmed respiratory viral illnesses, compared to previous years, primarily driven by RSV and influenza. A larger proportion of inpatients required Level 2 care during the current period. While more children have been admitted to CHEO during this time, to-date their overall LOSs are comparable to pre-pandemic periods.

SARS-CoV-2-related admissions were notably elevated in the months prior to the increase in RSV and influenza cases, and during the 2021/22 Winter period. While acute SARS-CoV-2 infection is not driving the current surge of relevant admissions, its impact on the immune system function of infants and children, and subsequent susceptibility to other pathogens, requires further investigation (8).

The progression of respiratory viral activity among children in our region is comparable to influenza activity recently experienced by Australia (9), with an earlier, steeper increase and higher volumes compared to pre-pandemic years. Their peak was followed by a sharp and sustained decrease in cases; the duration of our current viral season is, as of yet, undefined. In South Africa, the 2022 influenza season was more protracted, with percent positivity exceeding 10% for 25 weeks (3). European surveillance systems also reported earlier and increased RSV circulation, and median test positivity of 12.8% by Week 47 _(10)_. In Canada and the US, RSV test positivity during the current surveillance year peaked in early November and remains elevated at 7% as of December 10) (11). At CHEO, by December 10, our test positivity for influenza (26%; all samples have been Influenza A virus) was declining, RSV (9%) was declining, and SARS-CoV-2 (8%) was stable, reflecting ongoing high co-circulation of multiple respiratory viruses in the community (data from Eastern Ontario Regional Laboratory Association, not shown).

This descriptive analysis has numerous strengths. CHEO is the only pediatric health centre in Eastern Ontario and admits patients from western Quebec, Northern Ontario and Nunavut requiring tertiary-level care. Our dataset is comprehensive in capturing the burden of pediatric patients admitted with laboratory-confirmed respiratory viral disease over the five-year study period. Strategies to meet this increased volume have included creating additional PICU and inpatient medicine beds, changes to staffing models, and a recent provincial directive to transfer youth aged 14 years and up requiring intensive care to adult facilities as a decanting strategy (12,13).

We were able to examine changes in healthcare resource intensity by stratifying patient-level data according to the level of care received. As the lack of space in a conventional ICU setting has been a specific challenge during the current period, we recognized the limitations of comparing ICU admissions between surveillance years so instead, identified children who needed more resource-intense care, irrespective of their location in the hospital.

At a clinical level, our dataset demonstrates the evolution in care intensity related to high-flow nasal cannula oxygen therapy, introduced in Winter 2018/19 as standard of care for infants and children with moderate-to-severe respiratory distress (14). Given that this therapy is associated with an escalation of human and healthcare resource intensity to Level 2, we identified a pre-pandemic increase in the number of respiratory viral admissions requiring Level 2 but not Level 3 care. Further study is required to explain the higher proportion of Level 2 admissions in the current period.

As a single-centre study, the progression of pediatric respiratory viral admissions may not reflect the timing and trends in other jurisdictions, influenced by factors including pathogen transmissibility, community immunization coverage, pre-season SARS-CoV-2 activity, climate, and local and regional public health measures (15). Nonetheless, as the sole pediatric acute care facility for a large catchment area, we were able to provide population-level epidemiology, associated illness acuity and hospitalization resource needs which are generalizable to other midsize hospitals in public payer systems.

The study is limited to laboratory-confirmed respiratory viral infections and did not systematically capture post-infectious (including bacterial) complications that are known to occur following respiratory viral infections. Moreover, access to extended respiratory viral panel testing is restricted to critically ill children, high-risk patient populations and symptomatic neonates, so infections or coinfections with pathogens other than RSV, influenza and SARS-CoV-2 were missed, underestimating the true burden of the respiratory viral season in our cohort.

To our knowledge, this is the first study describing the impact of the current viral season on a Canadian pediatric hospital relative to previous surveillance periods. Our findings underscore the need for additional broad health human resources with expertise to care for critically ill infants and children, in addition to operational services to support front-line staff to provide safe and quality care.

We are repeating these analyses as the season progresses to better understand viral trends and levels of care; hospital leadership and policy makers can use these data to inform operational and human health resource planning for the current and future respiratory viral seasons.

## Data Availability

All data produced in the present work are contained in the manuscript

## Acknowledgements

The authors wish to thank the frontline clinicians and those who support them to provide safe and quality care, the CHEO Business Intelligence Team, and the CHEO Incident Management Team for their tireless efforts during this viral season.

