## Supplementary material for "Pediatric health system impact of an early respiratory viral season in Eastern Ontario, Canada: A descriptive analysis": Table S1

### Appendix - supplementary material

#### Supplementary 1: Patient length of stay by age and by detected respiratory virus, stratified by patient admission year

|  | **Patient admission year^a^** | | | | | | |
| --- | --- | --- | --- | --- | --- | --- | --- |
|  | **2017/18** | **2018/19** | **2019/20** | **2020/21** | **2021/22** | **2022/23^b^** | **Total** |
| **Patient LOS in days for admissions with a positive respiratory test^d^, by patient age at admission** |  |  |  |  |  |  |  |
| **≤5 months**  *median [Q1, Q3]* | 3.00 [2.00, 5.00] | 3.00 [2.00, 6.00] | 3.00 [2.00, 7.00] | 6.00 [2.75, 14.50] | 3.00 [2.00, 6.00] | 4.00 [2.00, 6.00] | 3.00 [2.00, 6.00] |
| ***p value*^c^** | *1.000* | *1.000* | *1.000* | *0.235* | *1.000* | *Ref.* |  |
| **6-11 months**  *median [Q1, Q3]* | 3.00 [2.00, 7.00] | 3.00 [2.00, 6.50] | 3.00 [2.00, 7.00] | 6.00 [4.25, 15.25] | 3.00 [1.00, 8.75] | 3.00 [2.00, 5.25] | 3.00 [2.00, 7.00] |
| ***p value*^c^** | *1.000* | *1.000* | *1.000* | *0.112* | *1.000* | *Ref.* |  |
| **12-23 months**  *median [Q1, Q3]* | 3.00 [2.00, 6.50] | 3.00 [2.00, 5.00] | 2.50 [2.00, 5.00] | 5.00 [2.50, 7.00] | 3.00 [2.00, 5.00] | 3.00 [2.00, 4.00] | 3.00 [2.00, 5.00] |
| ***p value*^c^** | *0.083* | *1.000* | *1.000* | *0.092* | *1.000* | *Ref.* |  |
| **2-4 years**  *median [Q1, Q3]* | 3.00 [2.00, 6.75] | 3.00 [2.00, 6.00] | 3.00 [2.00, 5.00] | 4.00 [3.00, 7.00] | 3.00 [2.00, 5.00] | 3.00 [2.00, 5.00] | 3.00 [2.00, 5.00] |
| ***p value*^c^** | *1.000* | *1.000* | *1.000* | *0.146* | *1.000* | *Ref.* |  |
| **5-9 years**  *median [Q1, Q3]* | 3.00 [2.00, 5.00] | 3.00 [2.00, 5.00] | 3.00 [2.00, 5.00] | 5.50 [3.25, 10.75] | 3.00 [2.00, 6.00] | 3.00 [2.00, 5.00] | 3.00 [2.00, 5.75] |
| ***p value*^c^** | *1.000* | *1.000* | *1.000* | *0.227* | *1.000* | *Ref.* |  |
| **≥10 years**  *median [Q1, Q3]* | 3.00 [2.00, 8.00] | 4.50 [2.25, 10.00] | 3.00 [2.00, 4.00] | 7.00 [1.50, 11.50] | 4.00 [2.00, 7.00] | 3.00 [2.00, 7.50] | 4.00 [2.00, 8.00] |
| ***p value*^c^** | *1.000* | *1.000* | *1.000* | *1.000* | *1.000* | *Ref.* |  |
| **Number of admissions with a positive respiratory test^d^**, **by virus** |  |  |  |  |  |  |  |
| **RSV** ^g^  *n (% per year group)* | 217 (47.4) | 327 (51.5) | 267 (53.4) | 0 (0) | 209 (35.5) | 345 (53.1) | 1365 (46.5) |
| **Influenza A or B**^e^  *n (% per year group)* | 99 (21.6) | 111 (17.5) | 125 (25) | 1 (1) | 13 (2.2) | 133 (20.5) | 482 (16.4) |
| **SARS-CoV-2**^e^  *n (% per year group)* | 0 (0) | 0 (0) | 1 (0.2) | 18 (17.5) | 199 (33.8) | 74 (11.4) | 292 (9.9) |
| **Other respiratory virus^ef^**  *n (% per year group)* | 154 (33.6) | 207 (32.6) | 128 (25.6) | 84 (81.6) | 191 (32.4) | 145 (22.3) | 909 (31) |

*^a^ Admission year grouping defined as starting on the 35^th^ week of one year and ending on the 34^th^ week of the following year*

*^b^ Time period is ongoing and incomplete*

*^c^* *Obtained via Kruskal-Wallis test (post hoc via Dunn’s Test of Multiple Comparisons with Bonferroni adjustment). Excludes Total column*

*^d^ Respiratory viruses were defined as one of the following: respiratory syncytial virus (RSV); Influenza A virus; Influenza B virus; SARS-CoV-2; or other viruses part of the extended respiratory viral panel (adenovirus; parainfluenza virus 1, 2, 3, and 4; human metapneumovirus, enterovirus/rhinovirus, seasonal coronaviruses). LOS and level of care were grouped by the peak period of the patients’ date of admission. The highest level of care was taken per patient.*

*^e^ Categories are not mutually exclusive*

*^f^ Other respiratory virus includes adenovirus; parainfluenza virus 1, 2, 3, and 4; human metapneumovirus, enterovirus/rhinovirus, seasonal coronaviruses*

*LOS, length of stay; Q1, first quartile; Q3, third quartile; RSV, respiratory syncytial virus*
